# Pooled sputum testing for the detection of pulmonary tuberculosis by Xpert^®^ MTB/RIF Ultra: a multi-site cross-sectional diagnostic evaluation study in Bangladesh

**DOI:** 10.64898/2026.03.19.26348771

**Authors:** S. M. Mazidur Rahman, Sohag Miah, Tanjina Rahman, Sabrina Choudhury, Noshin Nawer Ruhee, Senjuti Kabir, Mohammad Khaja Mafij Uddin, Shahriar Ahmed, Vibol Iem, Rachel L. Byrne, Ana I. Cubas Atienzar, Tushar Garg, Jacob Creswell, Tom Wingfield, Sayera Banu, Start4All Investigators

## Abstract

**Background:** Xpert^®^ MTB/RIF Ultra (Xpert Ultra) is a WHO recommended molecular test for rapid tuberculosis (TB) detection, but cartridge cost limits large-scale use of Xpert Ultra in high TB burden, resource-constrained countries. This study assessed the diagnostic performance of Xpert Ultra on pooled sputum for detection of pulmonary TB (PTB)in Bangladesh.

**Methods:** Between July 2024 and February 2025, adults with presumptive PTB were prospectively enrolled from primary and secondary healthcare facilities through facility-based case finding, and from urban slum communities through community-based case finding. Participants provided two sputum samples, one for individual and pooled Xpert Ultra testing, and another for culture. Pooled and individual Xpert Ultra results were compared to the microbiological reference standard of culture. Cost analysis was performed by comparing cartridge usage between individual and pooled Xpert Ultra testing.

**Results:** A total of 3043 individuals were tested individually and as part of 771 pooled samples. Compared with culture, the overall sensitivities of pooled and individual Xpert Ultra were 85.8% (95% CI: 79.8-90.6) and 89.2% (83.7-93.4), respectively, while specificities were 98.9% (98.4-99.2) and 98.1% (97.5-98.6). Pooled Xpert Ultra detected 100% of the high, medium, and low burden categories identified by individual testing, but showed lower detection for very low (81.8%) and trace (31.4%). Compared with individual testing, pooled testing reduced cartridge use and cost by 55.8%.

**Conclusions:** Pooled sputum testing with Xpert Ultra demonstrated high diagnostic performance similar to individual testing, while substantially reducing the cartridge costs. This approach offers a scalable molecular TB testing in resource-limited, high-burden countries such as Bangladesh.

## Introduction

Tuberculosis (TB) continues to be a major global health threat, particularly in low and middle income countries (LMICs) where diagnostic and treatment gaps persist[1]. Although being treatable and curable, in 2024,an estimated 10.7 million people developed TB globally, of whom 8.3 million were notified [1]. Bangladesh, one of the WHO high TB burden countries, reported 384,000 new TB cases in 2024, with an estimated 69,000 remaining undiagnosed[1]. The persistent diagnostic gap is due to multiple factors including limited access to healthcare services, diagnostic delay[2, 3], high out-of-pocket costs[4, 5], and inadequate healthcare infrastructure[2]. These challenges are particularly common in slum communities, where overcrowding and poor ventilation contribute to further TB transmission[6].

The Xpert^®^ MTB/RIF Ultra (Xpert Ultra) assay (Cepheid, Sunnyvale, California), first recommended by the WHO in 2017 as a first line TB diagnostic test, represents a significant advancement over its precursor, the Xpert^®^ MTB/RIF assay due to its improved sensitivity[7, 8]. However, the high cost of Xpert Ultra cartridges limits wide spread implementation in LMICs[9–12]. Consequently, National Tuberculosis Programme (NTP) often restricts the number of tests and placement of machines due to budget constraints[12]. In Bangladesh, only 52% of individuals diagnosed with TB in 2024 were tested with rapid molecular diagnostics, mostly with Xpert Ultra[1].

To increase the affordability of Xpert Ultra testing in lieu of price reductions, pooled sputum testing can be considered. Pooled sputum testing involves combining sputum specimens from multiple individuals into a single test[9, 13, 14]. If a pool test yields a positive result, all individual samples are retested to identify the positive sample(s). If the pool is negative, all samples in the pool are considered negative and reported as such. Evidence, mainly from single-country studies, has suggested that pooled testing can reduce cartridge use by 27%-49%, depending on TB prevalence in the target population, while maintaining high diagnostic sensitivity[13, 15, 16].

Previous studies have evaluated the performance of sputum pooling with Xpert Ultra, reporting positive percent agreement (PPA) compared to individual Xpert Ultra testing between 90% to 100%[9, 12, 17, 18]. A study in primary healthcare settings of Brazil reported pooled Xpert Ultra PPA and negative percent agreement (NPA) of 95% and 97.1%, respectively, achieving a 12.4% reduction in cartridge use[19]. In Nigeria, a study in secondary healthcare facilities reported PPA and NPA of 83.3% and 98.8%, respectively, with 46% cartridge savings[10]. Similarly, research in Laos People’s Democratic Republic demonstrated nearly complete agreement between pooled and individual tests[9].

Despite these promising findings, evidence on sputum pooling with Xpert Ultra remains limited across diverse healthcare facilities and slum communities in high TB burden regions in Southeast Asia, including Bangladesh. To date, no studies have used culture as a reference standard to provide complete diagnostic accuracy results. In the current study, we aimed to evaluate the performance of sputum pooling using Xpert Ultra in Bangladesh, stratified across primary and secondary level of healthcare facilities and urban slum community settings.

## Materials and Methods

### Study Settings and participant enrolment

This prospective cross-sectional diagnostic accuracy study was conducted between July 2024 and February 2025 under a Unitaid-funded multi-country project ‘Start4All’ (https://clinicaltrials.gov/study/NCT05845112). In Bangladesh, the study was carried out in two primary healthcare facilities (PHCFs) and two secondary healthcare facilities (SHCFs) in Rajshahi and Narsingdi districts, and urban slum communities (USCs) in Dhaka metropolitan city. In healthcare facilities, people with presumptive PTB were enrolled through a facility-based case finding (FBCF) approach, where individuals were screened using the WHO-recommended four-symptoms screen (W4SS)[20]. Individuals with at least two of the W4SS (cough, fever, weight loss and night sweats) or a cough of two weeks or more were eligible for inclusion. In contrast, participant enrollment in urban slum communities was conducted through a community-based case finding (CBCF) approach involving regular health campaigns in various slum areas of Dhaka city. Residents were invited to a designated location to assess their lung health condition using artificial intelligence (AI) enabled portable X-ray machine (MinXray equipped with Qure.ai software, version 3.2). Screening was performed with the W4SS criteria and AI-enabled portable X-ray machine with computer aided detection (CAD), and participants were enrolled if they had positive W4SS or a CAD score more than 0.3.

Individuals currently undergoing TB treatment (counting from third dose) or who had completed treatment within the previous six months were excluded. The study was conducted in accordance with the Standards for Reporting Diagnostic Accuracy (STARD) guideline[21]. The study was approved by the Research ethics committees of International Centre for Diarrhoeal Disease Research, Bangladesh (icddr,b), Liverpool School of Tropical Medicine, and WHO, and was performed in accordance with the Helsinki Declaration of 1964 and its later amendments. Written informed consent was obtained from all study participants before enrollment.

### Specimen collection and procedure

From each participant, two sputum samples (around 5 mL each) were collected. One sample was used for performing Xpert Ultra (both individual and pooled Xpert Ultra) at PHCFs and SHCFs, or nearby GeneXpert facilities for USCs. The second sputum sample was stored at 2-8 °C and transported to the Mycobacteriology laboratory of icddr,b for culture processing.

Xpert Ultra testing was performed according to the manufacturer’s standard operating procedures on sputum specimens, using a 1:2 sputum to sample reagent (SR) buffer ratio (Cepheid, Sunnyvale, California). Each sample was vortexed and incubated at room temperature for 15 minutes, with additional vortexing midway through incubation. Following incubation, 2 mL of the sputum-SR mixture was transferred into an Xpert Ultra cartridge, which was then loaded onto the GeneXpert platform for automated analysis. [8].

For pooled testing, remnant sputum-SR mixture from consecutive participants were typically combined in groups of four to create a single pooled sample, and tested on the same collection day within 4 hours if stored at room temperature or within 24 hours if stored at 2-8 °C. When the four samples were not available on the same day, and storage was not possible, pools were prepared with available three or two samples. A minimum of 1 mL of remnant sputum buffer mixture from each individual was combined to obtain a pooled volume of at least 2 mL. The pooled sample was then tested using an Xpert Ultra cartridge following the same procedure as for individual samples [9, 10] (**Figure 1**).

**Figure 1.**
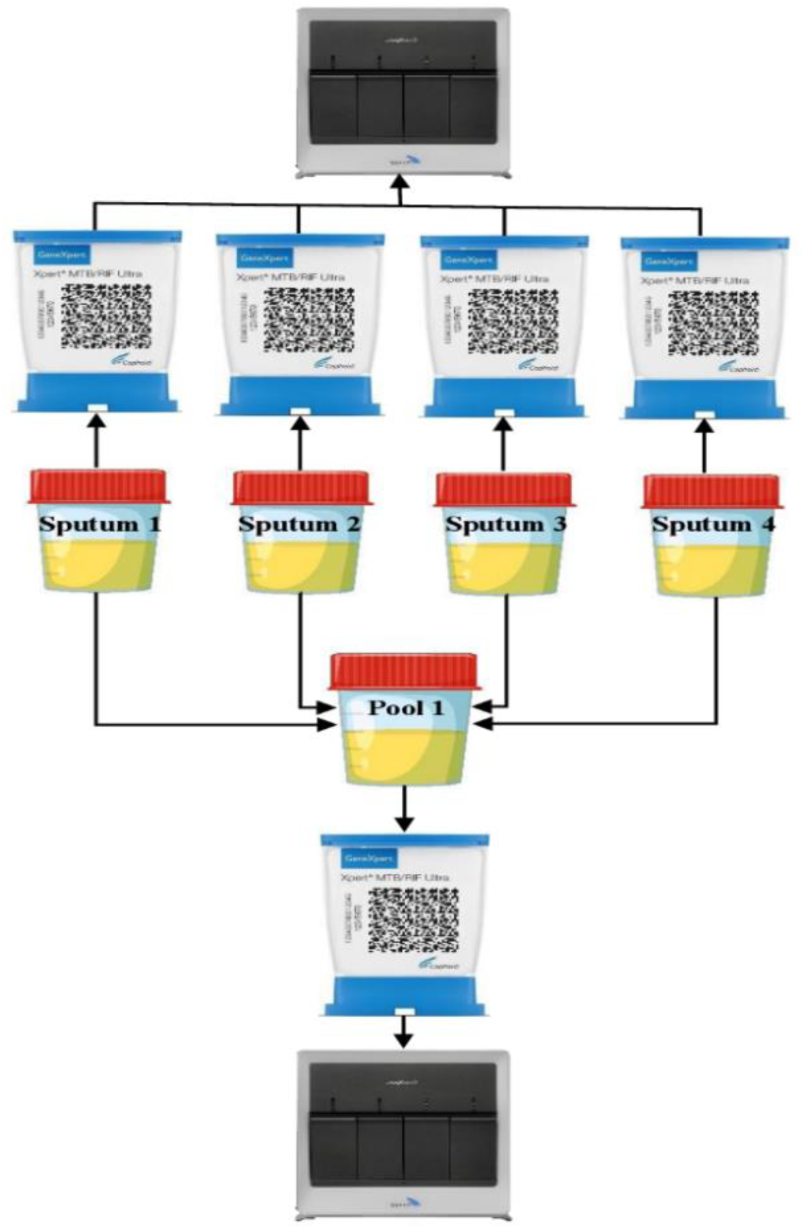
Workflow of individual and pooled Xpert Ultra. Individual sputum samples were processed with sample reagent (SR) ratio of 1:2. After that, 2 mL of the processed samples were loaded into the GeneXpert Ultra cartridge for testing. From the remaining of each of the processed sputum samples (approximately 4 sputum samples), ≥ 0.5 mL was transferred to a new cup mixing together to make a pool. Then from the pooled samples 2 mL of the processed sputum samples were transferred to a new GeneXpert Ultra cartridge for testing.

The second sputum sample was decontaminated and cultured in liquid media using BACTEC^TM^ MGIT^TM^ 960 system (BD Diagnostics Systems, USA) by following the procedure described previously[22, 23]. The culture growth was monitored continuously, and the machine generated results were recorded.

### Data Analysis

To evaluate the diagnostic performance, participants with valid results on pooled and individual Xpert Ultra, and culture were included. Sensitivity, specificity, positive predictive value (PPV), negative predictive value (NPV), overall accuracy, and kappa value (weighted) of pooled and individual Xpert Ultra were calculated with 95% confidence interval (CI) using exact binomial method at individual level against the results of culture. Positive percentage agreement (PPA) of pooled Xpert Ultra was determined against the results of individual Xpert Ultra. Passing-Bablok regression and Bland Altman analysis was used to assess the relationship of Ct values between the pooled and individual Xpert Ultra results across the multicopy IS6110-IS1081 target, and rpoB probes (rpoB1, rpoB2, rpoB3 and rpoB4). Cost analysis of pooled sputum testing was assessed by comparing the usage of cartridges between the pooled and individual Xpert Ultra testing strategies. As per GDF catalogue the cost of individual cartridge $7.97 was considered[24]. All statistical analyses were performed using STATA version 15.0 (StataCorp, College Station, TX, USA), IBM SPSS Statistics version 20.0 (IBM Corp., Armonk, NY, USA), and R version 4.4.2 (R Foundation for Statistical Computing, Vienna, Austria).

## Results

A total of 3050 individuals presumptive with PTB were enrolled, among whom results of Xpert Ultra on pooled and individual sputum samples were available for 3043 participants, and culture on individual sputum samples were available for 3034 participants. Overall study flowchart is shown in **Figure 2**.

**Figure 2.**
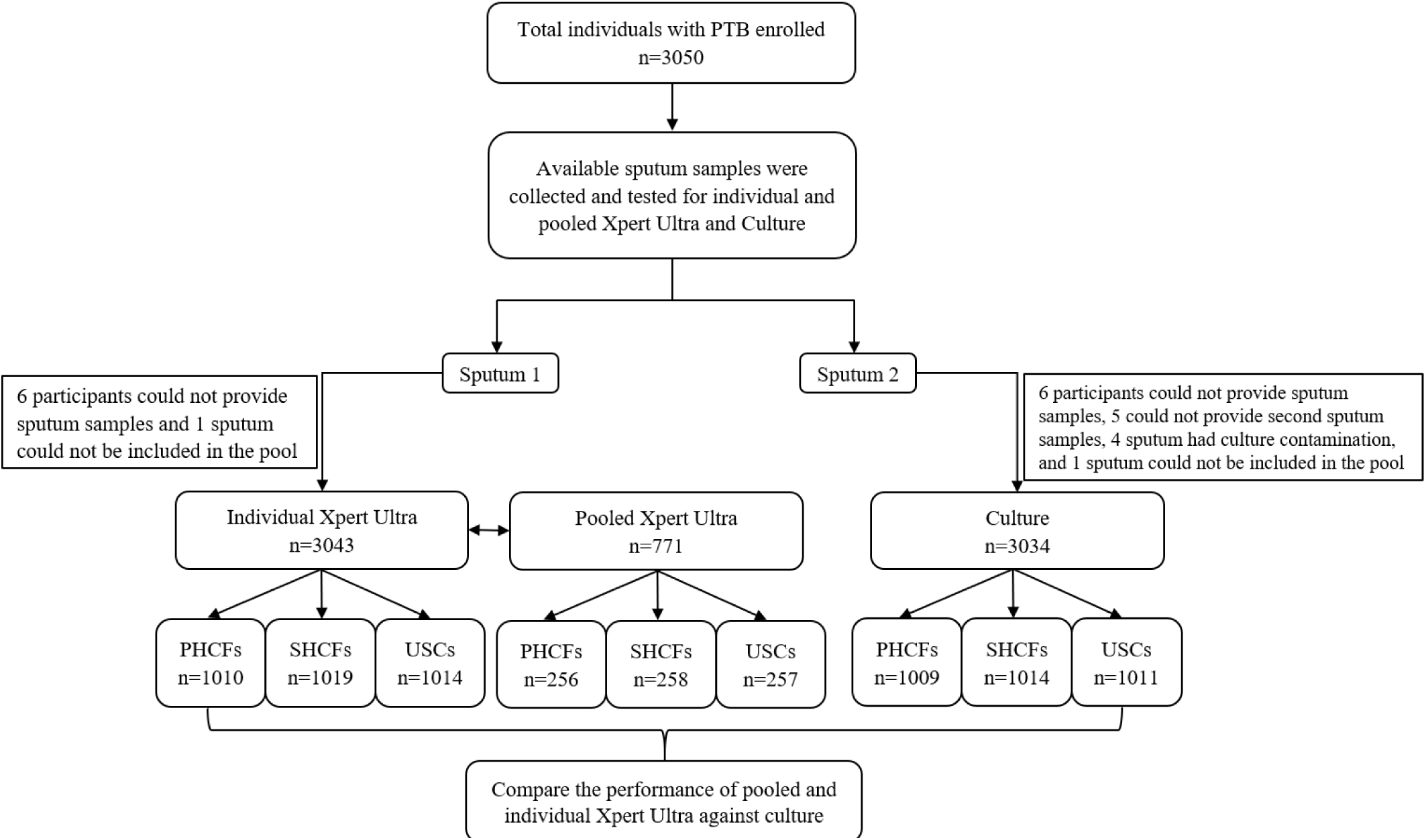
Study flowchart illustrating the enrolment of participants from primary healthcare facilities (PHCFs), secondary healthcare facilities (SHCFs), and urban slum communities (USCs) for the diagnosis of pulmonary tuberculosis (PTB). The collected sputum samples from participants were tested with Xpert MTB/RIF Ultra (Xpert Ultra) individually and by pooling. Each sputum sample was also cultured using BACTEC MGIT 960 system. Sputum samples with complete results of Xpert Ultra on individual and pooled testing as well as culture were included.

### Demographic and clinical characteristics of the enrolled participants

Of 3043 participants that had corresponding pooled and individual Xpert Ultra results, 1010, 1019, and 1014 were from PHCFs, SHCFs, and USCs, respectively. The majority of the enrolled participants were male (58.7%), and aged over 50 years (43.3%). Persistent cough lasting more than two weeks was the most common symptoms (90.2%), followed by fever (63.8%), weight loss (26.3%), and night sweat (21.5%). Most sputum samples collected from the participants were mucoid to mucopurulent (89.8%) (**Table 1**).

**Table 1.** Demographic and clinical characteristics of individuals with presumptive pulmonary TB enrolled from primary and secondary healthcare facilities and urban slum communities in Bangladesh.

| Characteristics | Total, n (%) | Passive Case Finding approach |  | Active Case Finding approach |
| --- | --- | --- | --- | --- |
|  |  | PHCFs, n (%) | SHCFs, n (%) | USCs, n (%) |
| **Participants** | 3043 | 1010 | 1019 | 1014 |
| **Sex** |  |  |  |  |
| Male | 1786 (58.7%) | 562 (55.6%) | 621 (60.9%) | 603 (59.5%) |
| Female | 1257 (41.3%) | 448 (44.4%) | 398 (39.1%) | 411 (40.5%) |
| **Age, years** |  |  |  |  |
| < 30 | 555 (18.2%) | 157 (15.5%) | 173 (17.0%) | 225 (22.2%) |
| 30-50 | 1170 (38.5%) | 383 (38.0%) | 355 (34.8%) | 432 (42.6%) |
| > 50 | 1318 (43.3%) | 470 (46.5%) | 491 (48.2%) | 357 (35.2%) |
| **Sputum quality** |  |  |  |  |
| Saliva | 292 (9.6%) | 173 (17.1%) | 51 (5.0%) | 68 (6.7%) |
| Mucoid | 2248 (73.9%) | 744 (73.7%) | 799 (78.4%) | 705 (69.5%) |
| Mucopurulent | 484 (15.9%) | 90 (8.9%) | 168 (16.5%) | 226 (22.3%) |
| Purulent | 19 (0.6%) | 3 (0.3%) | 1 (0.1%) | 15 (1.5%) |
| **Sputum blood** |  |  |  |  |
| Yes | 32 (1.1%) | 22 (2.2%) | 5 (0.5%) | 5 (0.5%) |
| No | 3011 (98.9%) | 988 (97.8%) | 1014 (99.5%) | 1009 (99.5%) |
| **WHO four symptom criteria** |  |  |  |  |
| Cough ($\geq 2$ weeks) | 2745 (90.2%) | 992 (98.2%) | 1010 (99.1%) | 743 (73.3%) |
| Fever | 1940 (63.8%) | 926 (91.7%) | 463 (45.4%) | 551 (54.3%) |
| Weight loss | 800 (26.3%) | 280 (27.7%) | 56 (5.5%) | 464 (45.8%) |
| Night sweat | 653 (21.5%) | 355 (35.1%) | 178 (17.5%) | 120 (11.8%) |

Abbreviations: PHCFs, primary healthcare facilities; SHCFs, secondary healthcare facilities; USCs, Urban slum communities

### Results of Xpert Ultra on individual and pooled sputum samples across different settings

A total of 3043 individual sputum specimens representing 771 pools were tested using the Xpert Ultra assay. The positivity rate of Xpert Ultra using individual testing was 7.0% (212/3043), with the highest detection rate in the USCs (9.6%, 97/1014) followed by SHCFs (6.1%, 62/1019) and PHCFs (5.3%, 53/1010). Among 212 individual Xpert Ultra positive sputum, the bacterial load with ‘High’, ‘Medium’, ‘Low’, and ‘Very Low’ category was 33.0%, 15.1%, 18.9%, and 12.3%, respectively. ‘Trace’ detected category among the individual Xpert Ultra positive results was 20.8% (**Table 2**).

**Table 2.**
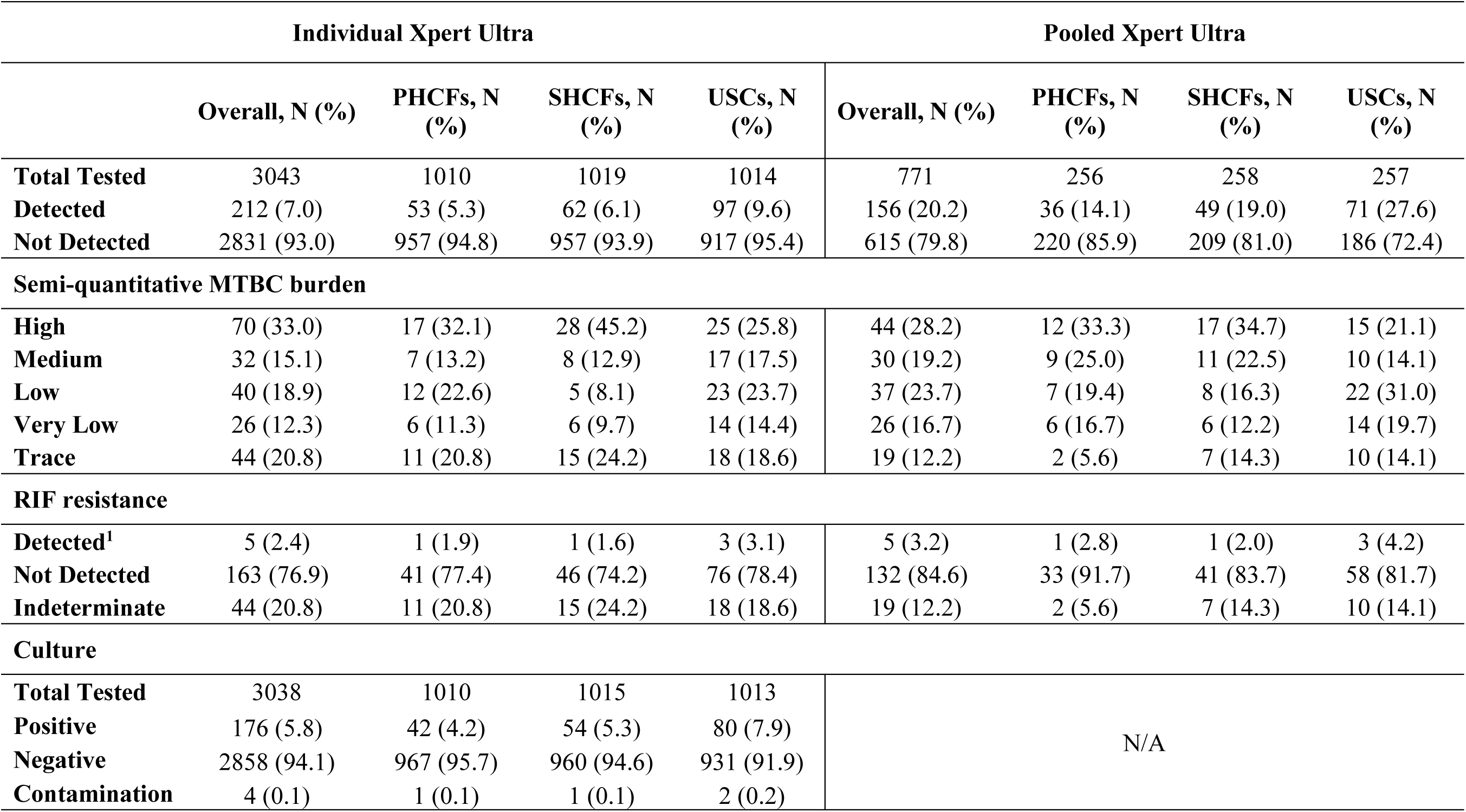
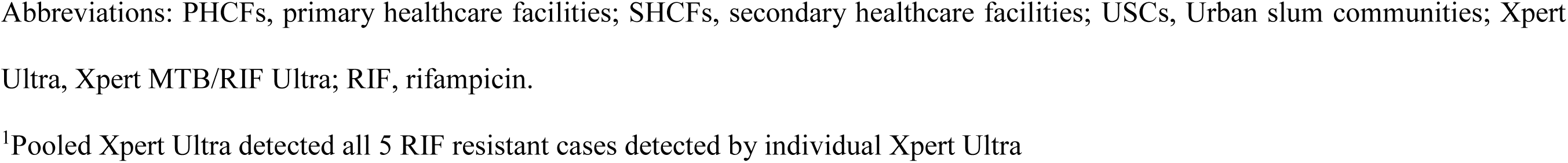
Results of Xpert Ultra on individual and pooled sputum samples, and culture on individual sputum samples across different settings.

|  | Individual Xpert Ultra |  |  |  | Pooled Xpert Ultra |  |  |  |
| --- | --- | --- | --- | --- | --- | --- | --- | --- |
|  | Overall, N (%) | PHCFs, N (%) | SHCFs, N (%) | USCs, N (%) | Overall, N (%) | PHCFs, N (%) | SHCFs, N (%) | USCs, N (%) |
| **Total Tested** | 3043 | 1010 | 1019 | 1014 | 771 | 256 | 258 | 257 |
| **Detected** | 212 (7.0) | 53 (5.3) | 62 (6.1) | 97 (9.6) | 156 (20.2) | 36 (14.1) | 49 (19.0) | 71 (27.6) |
| **Not Detected** | 2831 (93.0) | 957 (94.8) | 957 (93.9) | 917 (95.4) | 615 (79.8) | 220 (85.9) | 209 (81.0) | 186 (72.4) |
| **Semi-quantitative MTBC burden** |  |  |  |  |  |  |  |  |
| **High** | 70 (33.0) | 17 (32.1) | 28 (45.2) | 25 (25.8) | 44 (28.2) | 12 (33.3) | 17 (34.7) | 15 (21.1) |
| **Medium** | 32 (15.1) | 7 (13.2) | 8 (12.9) | 17 (17.5) | 30 (19.2) | 9 (25.0) | 11 (22.5) | 10 (14.1) |
| **Low** | 40 (18.9) | 12 (22.6) | 5 (8.1) | 23 (23.7) | 37 (23.7) | 7 (19.4) | 8 (16.3) | 22 (31.0) |
| **Very Low** | 26 (12.3) | 6 (11.3) | 6 (9.7) | 14 (14.4) | 26 (16.7) | 6 (16.7) | 6 (12.2) | 14 (19.7) |
| **Trace** | 44 (20.8) | 11 (20.8) | 15 (24.2) | 18 (18.6) | 19 (12.2) | 2 (5.6) | 7 (14.3) | 10 (14.1) |
| **RIF resistance** |  |  |  |  |  |  |  |  |
| **Detected<sup>1</sup>** | 5 (2.4) | 1 (1.9) | 1 (1.6) | 3 (3.1) | 5 (3.2) | 1 (2.8) | 1 (2.0) | 3 (4.2) |
| **Not Detected** | 163 (76.9) | 41 (77.4) | 46 (74.2) | 76 (78.4) | 132 (84.6) | 33 (91.7) | 41 (83.7) | 58 (81.7) |
| **Indeterminate** | 44 (20.8) | 11 (20.8) | 15 (24.2) | 18 (18.6) | 19 (12.2) | 2 (5.6) | 7 (14.3) | 10 (14.1) |
| **Culture** |  |  |  |  |  |  |  |  |
| **Total Tested** | 3038 | 1010 | 1015 | 1013 | N/A |  |  |  |
| **Positive** | 176 (5.8) | 42 (4.2) | 54 (5.3) | 80 (7.9) |  |  |  |  |
| **Negative** | 2858 (94.1) | 967 (95.7) | 960 (94.6) | 931 (91.9) |  |  |  |  |
| **Contamination** | 4 (0.1) | 1 (0.1) | 1 (0.1) | 2 (0.2) |  |  |  |  |

Abbreviations: PHCFs, primary healthcare facilities; SHCFs, secondary healthcare facilities; USCs, Urban slum communities; Xpert Ultra, Xpert MTB/RIF Ultra; RIF, rifampicin. <sup>1</sup>Pooled Xpert Ultra detected all 5 RIF resistant cases detected by individual Xpert Ultra

Xpert Ultra testing of 771 pooled sputum yielded an overall positivity rate of 20.2% (156/771), with highest in USCs (27.6%, 71/257) followed by SHCFs (19.0%, 49/258) and PHCFs (14.1%, 36/256). Among 156 positive pooled test results, ‘High’, ‘Medium’, ‘Low’, ‘Very Low’, and ‘Trace’ category were 28.2%, 19.2%, 23.7%, 16.7%, and 12.2%, respectively. Rifampicin (RIF) resistance was detected in 2.4% (5/212) positive individual sputum samples which was also detected by pooled Xpert Ultra. The overall positivity rate of culture was 5.8% (176/3038), with the highest rate was observed in USCs (7.9%, 80/1013) followed by 4.2% (42/1010) in PHCFs and 5.3% (54/1015) in SHCFs (**Table 2**).

Among the 771 pooled sputum samples, 95.8%, 3.0%, and 1.2% consisted of four, three, and two samples, respectively. Among 156 pooled Xpert Ultra positives, 83.3%, 15.4%, and 1.3%had one, two, and three individual positive samples, respectively. There was no Xpert Ultra positive pool sample with four individual positives (**Table 3**).

**Table 3.** Number of pools with 1, 2, 3 individual Xpert Ultra positive samples.

| Sample number<br>in a pool | All negative<br>(N=615)<br>n (%) | One positive<br>(N=130)<br>n (%) | Two positives<br>(N=24)<br>n (%) | Three positives<br>(N=2)<br>n (%) | Total number of pools<br>(N=771)<br>n (%) |
| --- | --- | --- | --- | --- | --- |
| **Four** | 587 (95.4%) | 126 (96.9%) | 24 (100.0%) | 2 (100.0%) | 739 (95.8%) |
| **Three** | 22 (3.6%) | 1 (0.8%) | 0 (0.0%) | 0 (0.0%) | 23 (3.0%) |
| **Two** | 6 (1.0%) | 3 (2.3%) | 0 (0.0%) | 0 (0.0%) | 9 (1.2%) |

Note: There were no four-sample pools that had four positive individual Xpert Ultra samples.

### Performance of Xpert Ultra on pooled sputum samples

Compared to culture, the overall sensitivity and specificity of Xpert Ultra on individual sputum was 89.2% (95% CI: 83.7-93.4) and 98.1% (95% CI: 97.5-98.6), respectively. Whereas the sensitivity and specificity of pooled Xpert Ultra was 85.8% (95% CI: 79.8-90.6) and 98.9% (95% CI: 98.4-99.2), respectively. The overall accuracy of individual and pooled Xpert Ultra with culture was 97.6% (95% CI: 97.0-98.1) (Kappa: 0.80) and 98.1% (95% CI: 97.6-98.6) (Kappa: 0.83), respectively. When stratified by healthcare facility, the sensitivity of Xpert Ultra on individual sputum was higher at 92.9% (95% CI: 80.5-98.5) in PHCFs compared to 85.2% (95% CI: 72.9-93.4) in SHCFs. For pooled Xpert Ultra, higher sensitivity was also observed at 88.1% (95% CI: 74.4-99.3) in PHCFs followed by 81.5% (95% CI: 68.6-90.8) in SHCFs. The sensitivities of individual and pooled Xpert Ultra were 90.0% (95% CI: 81.2-95.6) and 87.5% (95% CI: 78.2-93.8), respectively for USCs. Whereas, the specificities of both individual and pooled Xpert Ultra remained high ranging from 97.5 to 99.3% across different settings (**Table 4**).

**Table 4.** Performance of Individual and Pooled Xpert Ultra results compared to culture.

|  |  |  | Culture |  |  |  |  |  |  |  |
| --- | --- | --- | --- | --- | --- | --- | --- | --- | --- | --- |
|  |  |  | Positive | Negative | Sensitivity<br>(95% CI) | Specificity<br>(95% CI) | PPV<br>(95% CI) | NPV<br>(95% CI) | Accuracy<br>(95% CI) | k-value |
| Individual<br>Xpert Ultra | Overall<br>(n=3034) | Positive | 157 | 54 | 89.2 (83.7-93.4) | 98.1 (97.5-98.6) | 74.4 (69.0-79.2) | 99.3 (99.0-99.6) | 97.6 (97.0-98.1) | 0.80 |
|  |  | Negative | 19 | 2804 |  |  |  |  |  |  |
|  | PHCFs<br>(n=1009) | Positive | 39 | 14 | 92.9 (80.5-98.5) | 98.6 (97.6-99.2) | 73.6 (62.2-82.5) | 99.7 (99.1-99.9) | 98.3 (97.3-99.0) | 0.81 |
|  |  | Negative | 3 | 953 |  |  |  |  |  |  |
|  | SHCFs<br>(n=1014) | Positive | 46 | 16 | 85.2 (72.9-93.4) | 98.3 (97.3-99.0) | 74.2 (63.6-82.6) | 99.2 (98.4-99.6) | 97.6 (96.5-98.5) | 0.78 |
|  |  | Negative | 8 | 944 |  |  |  |  |  |  |
|  | USCs<br>(n=1011) | Positive | 72 | 24 | 90.0 (81.2-95.6) | 97.5 (96.3-98.4) | 75.0 (66.8-81.8) | 99.2 (98.4-99.6) | 97.0 (95.7-97.9) | 0.80 |
|  |  | Negative | 8 | 907 |  |  |  |  |  |  |
| Pooled Xpert<br>Ultra | Overall<br>(n=3034) | Positive | 151 | 32 | 85.8 (79.8-90.6) | 98.9 (98.4-99.2) | 82.5 (76.9-87.0) | 99.1 (98.7-99.4) | 98.1 (97.6-98.6) | 0.83 |
|  |  | Negative | 25 | 2826 |  |  |  |  |  |  |
|  | PHCFs<br>(n=1009) | Positive | 37 | 7 | 88.1 (74.4-99.3) | 99.3 (98.5-99.7) | 84.1 (71.5-91.8) | 99.5 (98.8-99.8) | 98.8 (97.9-99.4) | 0.85 |
|  |  | Negative | 5 | 960 |  |  |  |  |  |  |
|  | SHCFs<br>(n=1014) | Positive | 44 | 12 | 81.5 (68.6-90.8) | 98.8 (97.8-99.4) | 78.6 (67.3- 86.7) | 99.0 (98.2-99.4) | 97.8 (96.7-98.6) | 0.79 |
|  |  | Negative | 10 | 948 |  |  |  |  |  |  |
|  | USCs<br>(n=1011) | Positive | 70 | 13 | 87.5 (78.2-93.8) | 98.6 (97.6-99.3) | 84.3 (75.7-90.3) | 98.9 (98.1-99.4) | 97.7 (96.6-98.6) | 0.85 |
|  |  | Negative | 10 | 918 |  |  |  |  |  |  |

Abbreviations: PHCFs, primary healthcare facilities; SHCFs, secondary healthcare facilities; USCs, Urban slum communities; Xpert
Ultra, Xpert MTB/RIF Ultra; PPV, positive predictive value; NPV, negative predictive value; k, kappa

Compared to individual Xpert Ultra, the overall PPA of pooled Xpert Ultra testing was 86.8% (95% CI: 81.5-91.0), which ranged from 83.0% to 90.3% at different settings (**Supplementary Table 1**).

Among 3043 individuals tested, 44 had ‘Trace’ positive results on Xpert Ultra which were excluded to assess any changes in the pooled Xpert Ultra performance against culture. After exclusion, overall sensitivity of pooled Xpert Ultra improved to 88.2% (95% CI: 82.2-92.7), with highest sensitivity at 91.9% (95% CI: 78.1-98.3) in PHCFs (**Supplementary Table 2**). Whereas the specificity of pooled Xpert Ultra was around 99.0%. When compared against the individual Xpert Ultra, the overall PPA of pooled Xpert Ultra increased at 97.6% and across different healthcare settings (95.2-100%) (**Supplementary Table 3**).

### Semi-quantitative Xpert Ultra MTBC burden on pooled sputum samples

**Table 5** compares pooled and individual Xpert Ultra results stratified by bacterial burden among 158 MTBC-positive samples. The PPA of pooled Xpert Ultra was 100% when the bacterial load of the single individual positive sample within the pool was ‘High’ (52/52), ‘Medium’ (21/21), or ‘Low’ (28/28). However, substantial downgrading was observed among samples with ‘High’ grade, only 55.8% remained as ‘High’, while others shifted to lower burden categories. Similar patterns of downgrading were observed among samples with initial ‘Medium’ and ‘Low’ burdens except for one sample in each category that shifted to ‘High’. Among 35 individual samples with a ‘Trace’ result, only 11 were detected in the pool, resulting in a PPA of pooled Xpert Ultra only 31.4% (**Table 5**). Downgrading of bacterial burden in pooled sample was also evident through a strong linear relationship between pooled and individual Xpert Ultra Ct values, with pooled testing yielding progressively higher Ct values as individual Ct values increased (**Figure 3**). The differences in the Ct values between the pooled and individual Xpert Ultra are provided in supplementary materials (**Supplementary Figure 1 and Table 4**).

**Figure 3.**
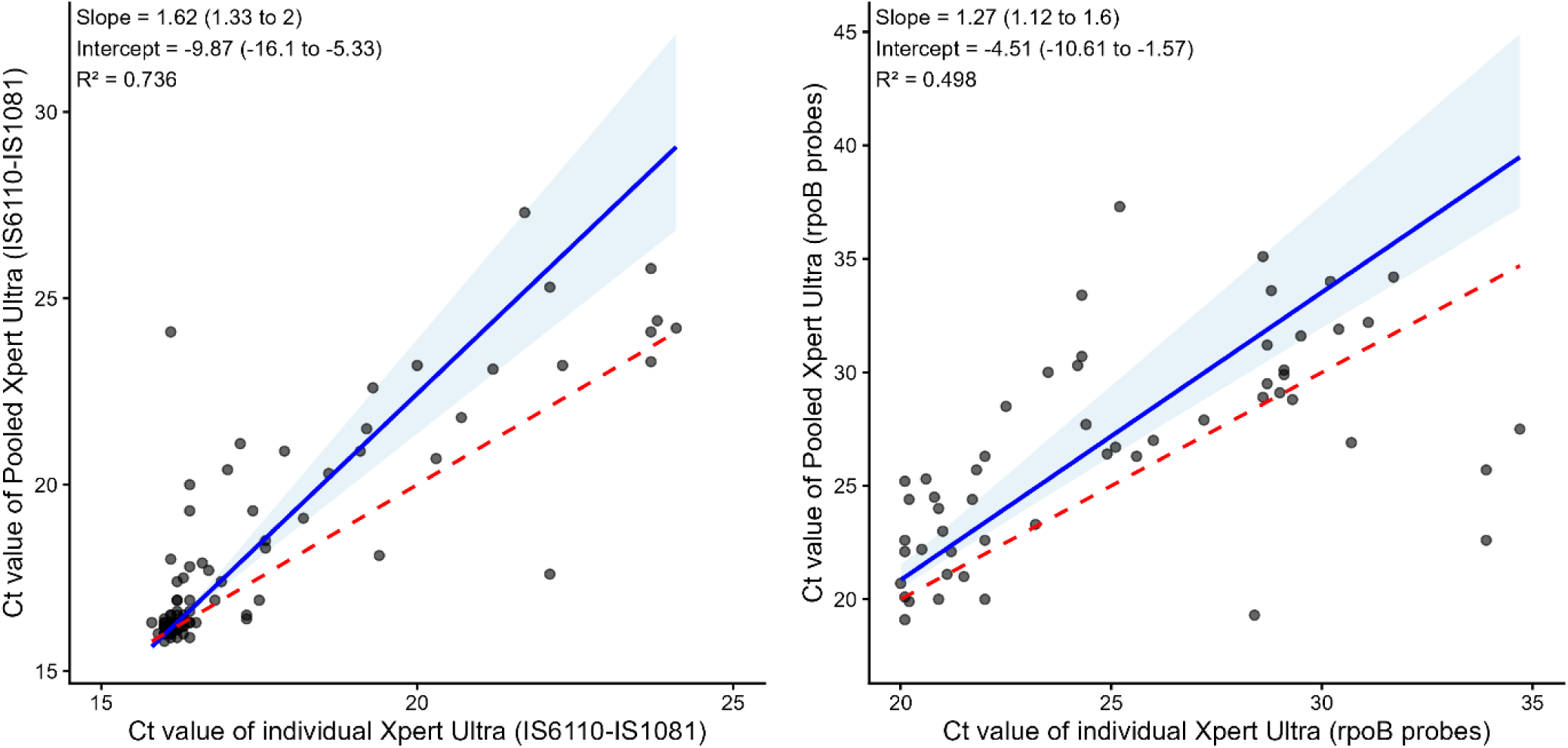
Passing-Bablok regression to evaluate the agreement between individual and pooled Xpert Ultra cycle threshold (Ct) values. The blue line is the regression line with 95% confidence intervals (shaded area), and the red dashed line is the line of equality. A strong positive association of the Ct values was observed between the pooled and individual Xpert Ultra for multicopy IS6110-IS1081 target (R² = 0.736) and rpoB probes (R² = 0.498), where pooled testing yielded progressively higher Ct values as individual Ct values increased. Pools with only one individual positive specimen were included.

**Table 5:**
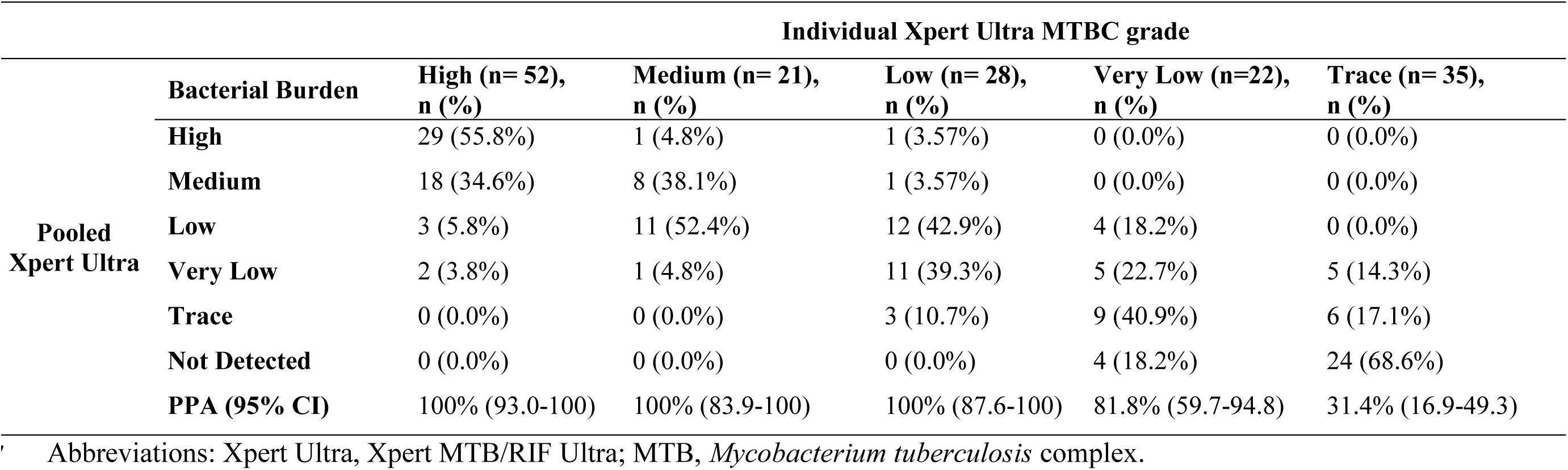
Distribution of semi-quantitative Xpert Ultra MTB burden in pooled and individual sputum samples (Only one positive sample included in the pool was considered)

### Cost saving by Xpert Ultra cartridge consumption for pooled and individual sputum samples testing

Among 771 pools, 156 tested positive, which required additional 617 individual Xpert Ultra testing for confirmation. Repeat Xpert Ultra testing due to error or invalid results was lower in pooled samples (3%) compared to individual samples (4.9%). Overall, a pooled testing approach would have reduced the total number of cartridges from 3192 (individual testing) to 1411, resulting in a reduction in total cost from $25,440.2 USD to $11,245.7 USD (at $7.97/test), corresponding to a cost saving of $14,194.6 USD (55.8%) for Xpert Ultra cartridges. The highest cost saving was observed in PHCFs (62.1%) followed by 56.2% in SHCFs and 48.7% in USCs (**Table 6**).

**Table 6.** Xpert Ultra cartridge related cost comparison between individual and pooled sputum samples testing.

| Parameters | Overall |  | PHCFs |  | SHCFs |  | USCs |  |
| --- | --- | --- | --- | --- | --- | --- | --- | --- |
|  | Individual Xpert Ultra | Pooled Xpert Ultra | Individual Xpert Ultra | Pooled Xpert Ultra | Individual Xpert Ultra | Pooled Xpert Ultra | Individual Xpert Ultra | Pooled Xpert Ultra |
| Total Samples | 3043 | – | 1010 | – | 1019 | – | 1014 | – |
| Total number of Pools | – | 771 | – | 256 | – | 258 | – | 257 |
| Positive Pools | – | 156 (20.2%) | – | 36 (14.1%) | – | 49 (19%) | – | 71 (27.6%) |
| Confirmatory Tests (Individuals in positive pools) | – | 617 | – | 144 | – | 196 | – | 280 |
| Repeat tests (Error/Invalid) | 149 (4.9%) | 23 (3%) | 75 (7.4%) | 11 (4.3%) | 40 (3.9%) | 10 (3.9%) | 34 (3.4%) | 2 (0.8%) |
| Total Tests Performed | 3192 | 1411 | 1085 | 411 | 1059 | 464 | 1048 | 537 |
| Total Cost (USD @ **\$7.97/test**) | \$25,440.2 | \$11,245.7 | \$8,647.5 | \$3,275.7 | \$8,440.2 | \$3,698.1 | \$8,352.6 | \$4,279.9 |
| Total Cost Saved | – | **\$14,194.6** | – | **\$5,371.8** | – | **\$4,742.2** | – | **\$4,072.7** |
| **% Cost Savings** | – | **55.8%** | – | **62.1%** | – | **56.2%** | – | **48.7%** |

Abbreviations: PHCFs, primary healthcare facilities; SHCFs, secondary healthcare facilities; USCs, Urban slum communities; Xpert
Ultra, Xpert MTB/RIF Ultra

## Discussion

This study provides the first evaluation of a sputum pooling strategy for the diagnosis of PTB in Bangladesh, a high TB-burden country where Xpert Ultra is routinely used for microbiological confirmation. We present both the diagnostic performance (sensitivity and specificity) and the potential cartridge and related monetary savings of pooled Xpert Ultra testing across diverse healthcare facilities and densely populated slums. Using culture as the reference standard, the overall sensitivity of pooled Xpert Ultra was 85.8%, similar to individual Xpert Ultra (89.2%), whereas the specificity remained high (>98%) for both pooled and individual Xpert Ultra. The modest reduction in sensitivity (3.4%) is likely attributable to dilution effects inherent to pooling, particularly for samples containing a low burden of MTBC-mostly the ‘Trace’ category, which tended to become negative when combined with other samples. This was further supported by our stratified analysis when ‘Trace’ results were excluded from the main evaluation, the sensitivity of pooled Xpert Ultra increased to 88.2% compared with culture, and PPA increased to 97.6% when compared with individual Xpert Ultra results. Similar observation was reported by Santos et al. (2023) in Brazil [19], who also noted improved accuracy (95.3%) when individual ‘Trace’ results were reclassified as negative.

Our findings are consistent with previous evaluations from other countries, which reported pooled Xpert Ultra PPA ranging from 83% to 100% and NPA from 97% to 100% when compared with individual Xpert Ultra testing [9, 10, 12, 15, 19]. An important aspect of our study is that we calculated the sensitivity of the pooled Xpert Ultra testing first time at individual level using MGIT culture as the gold standard, providing a more stringent assessment of diagnostic performance.

Across all the settings, pooled Xpert Ultra testing consistently yielded over 98% specificity This highlights the reliability of pooled testing to accurately identify people without TB and minimize false positive results, which is crucial for efficient resource utilization in high-burden, resource-limited contexts. Whereas, variation was observed in sensitivity across all settings with PHCFs had the highest sensitivity (88.1%). However, sensitivity improved across all sites when individual “Trace” results were excluded from the main evaluation, with PHCFs achieving highest sensitivity (91.9%). This improvement reveals how low bacillary load, particularly trace-positive samples, affects pooled testing’s diagnostic performance.

The semi-quantitative MTBC grade analysis revealed a consistent downward shift in bacillary load in pooled results, supported by higher Ct values on regression analysis. While pooled Xpert Ultra detected all ‘High’, ‘Medium’, and ‘Low’ burden samples (100%), most were downgraded to lower burden categories. Importantly, pooled strategy failed to reliably detect ‘Trace’-only positive samples, with PPA falling to 31.4%, consistent with previous studies [10, 19]. This has important clinical implications, as trace results may represent early or paucibacillary disease, where sensitivity is inherently limited [25]. In such population, particularly people living with HIV, hospitalized or critically ill patients, and those with prior history of TB, pooled testing may be suboptimal, and individual testing should be prioritized. Recent evidence from Uganda suggests that individuals with ‘Trace’ positive results and atypical chest X-ray findings have a higher risk of progression to active TB[26]. Therefore, individuals with abnormal chest X-ray or higher CAD scores who may be more likely to have paucibacillary TB could be considered for individual Xpert Ultra testing to minimize the risk of missed diagnosis. Further studies are needed to guide optimal use of pooled testing in population at higher risk of paucibacillary TB.

Our study demonstrates potential cost savings associated with pooled sputum testing using the Xpert Ultra assay compared to individual testing. Overall, using a pooled Xpert Ultra testing strategy would have reduced cartridge consumption by more than half, resulting in a 55.8% reduction in total costs. The highest cartridge-related cost savings were observed at PHCFs (62.1%), followed by SHCFs (56.2%) and USCs (48.7%). These findings are consistent with previous reports highlighting the economic benefits of pooled Xpert Ultra testing strategies, particularly in high-burden, resource-limited settings where Xpert testing capacity exists [11, 12]. Notably, pool testing was also associated with fewer repeat tests due to errors or invalid results compared to individual Xpert Ultra testing, indicating improved operational efficiency. Evidence from other countries and settings similarly shows that pooling can conserve resources while maintaining high diagnostic accuracy [9, 10, 12–14, 17, 29].

In Bangladesh, where cartridge supply remains constrained and many facilities continue to rely on microscopy, reflect by only 52% of diagnosed TB patients were tested with rapid molecular diagnostics in 2024, mostly with Xpert Ultra [1]. Under such circumstances, our findings highlight the strong potential of the pooled Xpert Ultra testing to maximize the use of limited cartridges and expand testing capacity for TB detection. This approach could be scaled up for both CBCF and FBCF approaches, as demonstrated in our study through substantial cost savings. However, pooled Xpert Ultra testing may provide greater benefit in CBCF as community screening involves large numbers of individuals and therefore maximizes the reduction in cartridge use and overall costs. By enabling testing of more people with the same resources pooled Xpert Ultra would facilitates earlier detection of individuals with TB who might not otherwise present to healthcare facilities.

Our study had some important limitations. First, individual and pooled Xpert Ultra testing was performed on the first sputum sample, whereas culture was performed on the second sputum collected 30 minutes later due to difficulty in splitting sputum specimens at lower levels of care and / or insufficient single sputum quantities. Nonetheless, the overall accuracy between pooled Xpert Ultra and culture exceeded over 97.6%, suggesting that using different samples had minimal effect on performance. Second, the cost analysis included only cartridge consumption, and did not account for additional expenses such as labor, staff training, infrastructure or transportation. Despite these limitations, the strengths of the current study are having a large sample size, inclusion of diverse healthcare and community settings, and the use of culture as the comparator for evaluating the performance of pooled Xpert Ultra. Importantly, the implementation cost of sputum pooling is minimal, as the approach does not require additional personnel or major workflow changes, and only minimal training is needed. This supports the feasibility of pooled testing at a programmatic level across the country. However, future research should focus on optimizing pooling strategies by integrating low-cost screening tools such as AI-enabled portable chest X-ray. For example, individuals with higher CAD scores could be prioritize for individual testing, while those with intermediate scores (regardless of symptoms) and low scores with symptoms could be grouped for pooling. Such targeted pooling could further enhance cartridge saving, expand testing coverage, and improve early detection of TB.

In conclusion, the performance of pooled Xpert Ultra was similar to that of individual Xpert Ultra in detecting PTB across both community and healthcare facilities in Bangladesh. Overall, pooled Xpert Ultra testing reduced cartridge consumption by more than half compared to individual testing, providing substantial savings in cost and resources. Although a minimal loss in sensitivity was observed, the overall benefit of testing large number of people with presumptive TB at lower cost makes pooled Xpert Ultra a compelling approach for programmatic adoption. Implementing this approach would particularly be helpful for high TB burden countries like Bangladesh to maintain uninterrupted testing during periods of financial constraint or supply shortages so as to reach all presumptive TB under molecular testing.

## Funding

The Start4All consortium and its work is funded by UNITAID, Grant number: 2022-50-START-4-ALL. Country specific grant number is: RPBS 05267,WR9. Views and opinions of authors expressed herein do not necessarily reflect the views of funder. The funder of this study had no role in the in the study design, data collection, analysis and interpretation, preparation of the manuscript, or decision to publication.

## Data Availability

All data produced in the present study are available upon reasonable request to the authors

## Acknowledgements

This study was conducted with the support of UNITAID. icddr,b acknowledges with gratitude the commitment of UNITAID to its research efforts. icddr,b is also grateful to the Governments of Bangladesh and Canada for providing core/unrestricted support. The authors acknowledge the national tuberculosis control program of Bangladesh for allowing us to conduct this research. The authors also thank the authorities and physicians of participating hospitals to permit us in conducting research activities at their healthcare facilities.

## Author contributions

J. C., T. W., and S. B.: conceptualization; S. M. M. R., S. M., N. N. R., S. C., M. K. M. U., T. G., V. I., R. L. B., and A. I. C. A.: methodology; S. M. M. R., S. C., S. K., V. I., and R. L. B.: validation; S. M. M. R., S. M., T. R., J. C., T. W., and V. I.: formal analysis; S. M. M. R., S. K., S. C., S.A. N. N. R., M. K. M. U., and S. B.: investigation; S. B., S. M. M. R., and T. W.: resources; S. M. M. R., S. M. and T. R.: data curation; S. M. M. R.: writing—original draft; all authors: writing—review and editing; S. M. M. R., S. C., S. A., and S.B.: supervision; S. M. M. R., and S. B.: administration; J. C., T. W., and S. B.: funding acquisition. All authors have read and agreed to the published version of the manuscript.

## Start4All investigators

Tom Wingfield (Centre for Tuberculosis Research & Departments of International Public Health and Clinical Sciences, Liverpool School of Tropical Medicine, UK. Tropical and Infectious Disease Unit, Liverpool University Hospitals NHS Foundation Trust, Liverpool, UK,); Jacob Creswell (Stop TB Partnership, Geneva, Switzerland,); Augustine Choko (Malawi Liverpool Wellcome Programme, Department of International Public Health, Liverpool School of Tropical Medicine, Blantyre, Malawi,); Melissa Sander (CHPR, Bamenda, Cameroon,); Sayera Banu (icddr,b, Dhaka, Bangladesh,); Victor Santana Santos (Federal University of Sergipe, Lagarto, Brazil,); Stephen John (Janna Health Foundation, Yola, Nigeria,); John Bimba (Zankli Research Centre, Bingham University, Karu, Nigeria,); Steve Otieno Wandiga (Kenya Medical Research Institute, Kisumu, Kenya,); Luan Nguyen Quang Vo (Friends for International TB Relief, Hanoi, Vietnam,); Andrew Codlin (Friends for International TB Relief, Ho Chi Minh City, Vietnam,).

## Conflicts of interest

The authors declare no conflicts of interests.

## Data sharing statement

The data underlying this article will be shared on reasonable request to the corresponding authors.

## Notes

### Competing Interest Statement

The authors have declared no competing interest.

### Author Declarations

Ethics committee of International Centre for Diarrhoeal Disease Research, Bangladesh (icddr,b), Liverpool School of Tropical Medicine (LSTM), and WHO gave ethical approval for this work

