## Supplemental file for "Pooled sputum testing for the detection of pulmonary tuberculosis by Xpert^®^ MTB/RIF Ultra: a multi-site cross-sectional diagnostic evaluation study in Bangladesh"

**Supplementary Table 1.** Performance of pooled Xpert Ultra compared to the results of individual Xpert Ultra

| **Pooled Xpert Ultra** |  | |  | | | | **Individual Xpert Ultra** | | |
| --- | --- | --- | --- | --- | --- | --- | --- | --- | --- |
|  |  | **True Positive (TP)** | | **False Negative (FN)** | | | | **True Negative (TN)** | **PPA (95% CI)** |
|  | **Overall (n=3043)** | 184 | | | 28 | 2831 | | | 86.8 (81.5-91.0) |
|  | **PHCFs (n=1010)** | 44 | | | 9 | 956 | | | 83.0 (70.2-91.9) |
|  | **SHCFs (n=1019)** | 56 | | | 6 | 957 | | | 90.3 (80.1-96.4) |
|  | **USCs (n=1014)** | 84 | | | 13 | 921 | | | 86.6 (78.2-92.7) |

Abbreviations: PHCFs, primary healthcare facilities; SHCFs, secondary healthcare facilities; USCs, Urban slum communities; Xpert Ultra, Xpert MTB/RIF Ultra; PPA, positive percent agreement

**Supplementary Table 2.** Performance of pooled Xpert Ultra compared to results of culture excluding the ‘Trace’ positive individuals

| **Pooled Xpert Ultra** |  |  | **Culture** | | | | | | |  |
| --- | --- | --- | --- | --- | --- | --- | --- | --- | --- | --- |
|  |  |  | **Positive** | **Negative** | **Sensitivity**  **(95% CI)** | **Specificity**  **(95% CI)** | **PPV**  **(95% CI)** | **NPV**  **(95% CI)** | **Agreement**  **(95% CI)** | ***k*-value** |
|  | **Overall (n=2990)** | **Positive** | 142 | 21 | 88.2 (82.2-92.7) | 99.3 (98.9-99.5) | 87.1 (81.5-91.2) | 99.3 (99.0-99.6) | 98.7 (98.2-99.0) | 0.87 |
|  |  | **Negative** | 19 | 2808 |  |  |  |  |  |  |
|  | **PHCFs (n=998)** | **Positive** | 34 | 6 | 91.9 (78.1-98.3) | 99.4 (98.7-99.8) | 85.0 (71.7-92.7) | 99.7 (99.1-99.9) | 99.1 (98.3-99.6) | 0.87 |
|  |  | **Negative** | 3 | 955 |  |  |  |  |  |  |
|  | **SHCFs (n=999)** | **Positive** | 41 | 6 | 83.7 (70.3-92.7) | 99.4 (98.6-99.8) | 87.2 (75.3-93.9) | 99.2 (98.4-99.6) | 98.6 (97.7-99.2) | 0.85 |
|  |  | **Negative** | 8 | 944 |  |  |  |  |  |  |
|  | **USCs (n=993)** | **Positive** | 67 | 9 | 89.3 (80.1-95.3) | 99.0 (98.2-99.6) | 88.2 (79.5-93.5) | 99.1 (98.3-99.5) | 98.3 (97.3-99.0) | 0.88 |
|  |  | **Negative** | 8 | 909 |  |  |  |  |  |  |

Abbreviations: PHCFs, primary healthcare facilities; SHCFs, secondary healthcare facilities; USCs, Urban slum communities; Xpert Ultra, Xpert MTB/RIF Ultra; PPV, positive predictive value; NPV, negative predictive value; k, kappa

**Supplementary Table 3.** Performance of pooled Xpert Ultra compared to the results of individual Xpert Ultra excluding the ‘Trace’ positive individuals

| **Pooled Xpert Ultra** |  | |  | | **Individual Xpert Ultra** | | |
| --- | --- | --- | --- | --- | --- | --- | --- |
|  |  | **True Positive (TP)** | | **False Negative (FN)** | | **True Negative (TN)** | **PPA (95% CI)** |
|  | **Overall (n=2999)** | 164 | | 4 | | 2831 | 97.6 (94.0-99.4) |
|  | **PHCFs (n=999)** | 40 | | 2 | | 957 | 95.2 (83.8-99.4) |
|  | **SHCFs (n=1004)** | 47 | | 0 | | 957 | 100.0 (92.5-100.0) |
|  | **USCs (n=996)** | 77 | | 2 | | 917 | 97.5 (91.2-99.7) |

Abbreviations: PHCFs, primary healthcare facilities; SHCFs, secondary healthcare facilities; USCs, Urban slum communities; Xpert Ultra, Xpert MTB/RIF Ultra; PPA, positive percent agreement

**Supplementary Table 4:** Median cycle threshold values between individual and pooled Xpert Ultra

|  | **Individual Xpert Ultra, n=107** | **Pooled Xpert Ultra, n=107** |  |
| --- | --- | --- | --- |
| **Variables** | **Ct Median (IQR)** | **Ct Median (IQR)** | **ΔCt, Median** |
| IS1081/IS6110 | 16.2 (16.1 - 17.2) | 16.3 (16.2 - 18.1) | 0.1 |
| rpoB1 | 18.9 (17.7 - 23.1) | 19.8 (18.5 - 25.8) | 0.9 |
| rpoB2 | 18.8 (17.6 - 22.8) | 19.6 (18.3 - 25.4) | 0.8 |
| rpoB3 | 20.2 (18.8 - 24.5) | 21.5 (19.7 - 27.4) | 1.3 |
| rpoB4 | 22.2 (21.0 - 26.9) | 23.5 (21.8 - 29.7) | 1.3 |

Abbreviations: Ct, Cycle threshold; IQR, Interquartile range; ΔCt, Median = (difference between pooled and individual median Ct values).

Difference between the pooled and individual Xpert Ultra median Ct value for IS6110-IS1081 probe was 0.1, whereas for the rpoB probes (rpoB1, rpoB2, rpoB3, and rpoB4) ranged from 0.8 to 1.3 cycles.

**Supplementary Figure 1**

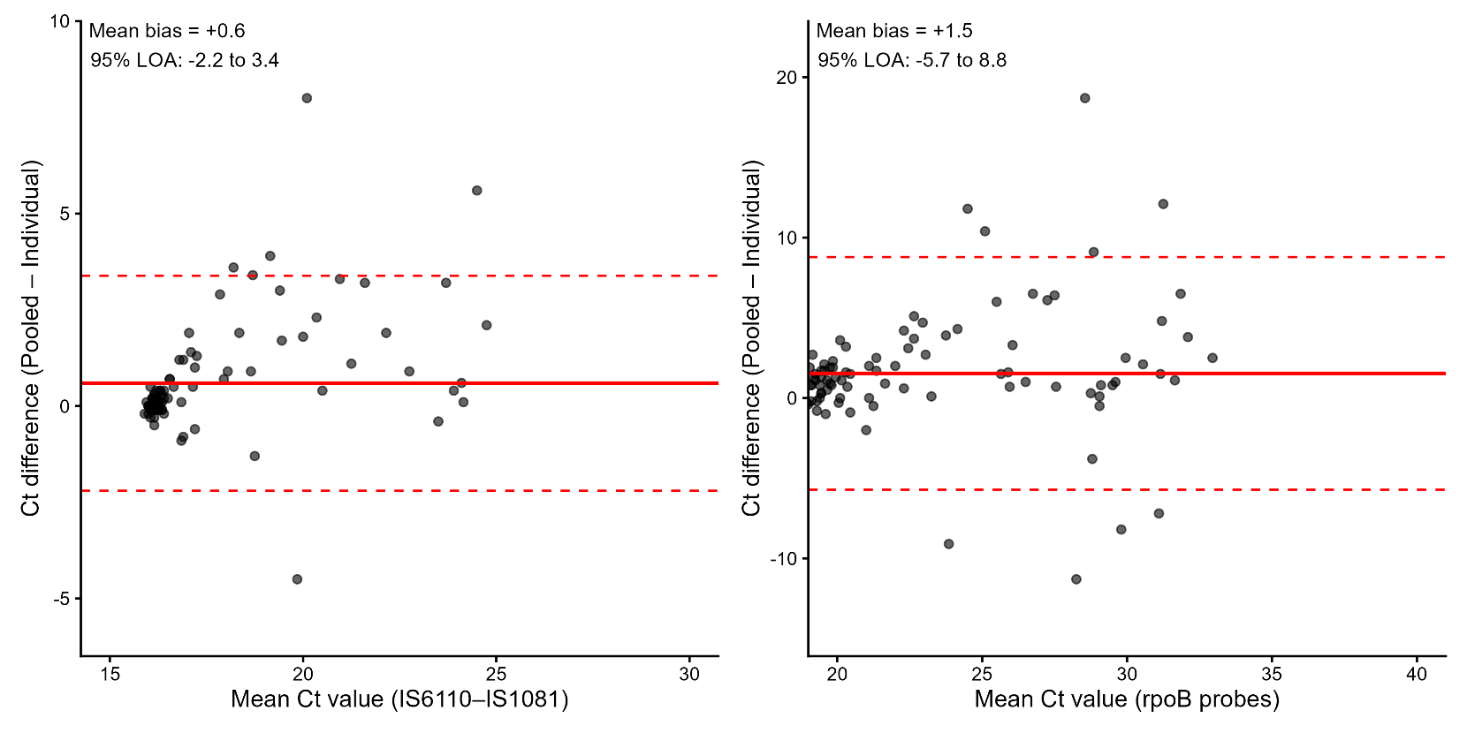

**Supplementary Figure 1:** Bland-Altman plots for assessing agreement between pooled and individual Xpert Ultra Ct values of the different genetic targets. The solid red line is the mean bias that indicates the systematic change in Ct values between the individual and pooled Xpert Ultra and the dashed line indicates the 95% limits of agreement (LOA). For IS6110-IS1081 target, the mean bias was +0.6 cycle (95% LOA, -2.2 to +3.4) and for rpoB probes, the mean bias was +1.5 cycles (95% LOA, -5.7 to +8.8), indicating a small increase in Ct values of the pooled Xpert Ultra when compared with individual Xpert Ultra. Pools with only one individual positive specimen were included.
